# Segmentation stability of human head and neck cancer medical images for radiotherapy applications under de-identification conditions: benchmarking data sharing and artificial intelligence use-cases

**DOI:** 10.1101/2022.01.22.22269695

**Authors:** Jaakko Sahlsten, Kareem A. Wahid, Enrico Glerean, Joel Jaskari, Mohamed A. Naser, Renjie He, Benjamin H. Kann, Antti Mäkitie, Clifton D. Fuller, Kimmo Kaski

## Abstract

**Background:** Demand for head and neck cancer (HNC) radiotherapy data in algorithmic development has prompted increased image dataset sharing. Medical images must comply with data protection requirements so that re-use is enabled without disclosing patient identifiers. Defacing, i.e., the removal of facial features from images, is often considered a reasonable compromise between data protection and re-usability for neuroimaging data. While defacing tools have been developed by the neuroimaging community, their acceptability for radiotherapy applications have not been explored. Therefore, this study systematically investigated the impact of available defacing algorithms on HNC organs at risk (OARs).

**Methods:** A publicly available dataset of magnetic resonance imaging scans for 55 HNC patients with eight segmented OARs (bilateral submandibular glands, parotid glands, level II neck lymph nodes, level III neck lymph nodes) was utilized. Eight publicly available defacing algorithms were investigated: afni_refacer, DeepDefacer, defacer, fsl_deface, mask_face, mri_deface, pydeface, and quickshear. Using a subset of scans where defacing succeeded (N=29), a 5-fold cross-validation 3D U-net based OAR auto-segmentation model was utilized to perform two main experiments: 1.) comparing original and defaced data for training when evaluated on original data; 2.) using original data for training and comparing the model evaluation on original and defaced data. Models were primarily assessed using the Dice similarity coefficient (DSC).

**Results:** Most defacing methods were unable to produce any usable images for evaluation, while mask_face, fsl_deface, and pydeface were unable to remove the face for 29%, 18%, and 24% of subjects, respectively. When using the original data for evaluation, the composite OAR DSC was statistically higher (p ≤ 0.05) for the model trained with the original data with a DSC of 0.760 compared to the mask_face, fsl_deface, and pydeface models with DSCs of 0.742, 0.736, and 0.449, respectively. Moreover, the model trained with original data had decreased performance (p ≤ 0.05) when evaluated on the defaced data with DSCs of 0.673, 0.693, and 0.406 for mask_face, fsl_deface, and pydeface, respectively.

**Conclusion:** Defacing algorithms may have a significant impact on HNC OAR auto-segmentation model training and testing. This work highlights the need for further development of HNC-specific image anonymization methods.

## Introduction

The landscape of data democratization is rapidly changing. The rise of open science practices, inspired by coalitions such as the Center for Open Science (1), and the FAIR (Findable, Accessible, Interoperable, and Reusable) guiding principles (2), has spurred interest in public data sharing. Subsequently, the medical imaging community has increasingly adopted these practices through initiatives such as The Cancer Imaging Archive (3). Given the appropriate removal of protected health information through anonymization techniques, public repositories have democratized the access to medical imaging data such that the world at large can now help develop algorithmic approaches to improve clinical decision-making. Among the medical professions seeking to leverage these large datasets, radiation oncology has the potential to vastly benefit from these open science practices (4). Imaging is crucial to radiotherapy workflows, particularly for organ at risk (OAR) and tumor segmentation (5,6). Moreover, in recent years public data competitions, such as the Head and Neck Tumor Segmentation and Outcome Prediction in positron emission tomography/computed tomography (PET/CT) Images (HECKTOR) challenge (7–9), have been targeted to improve the radiotherapy workflow. However, there is a particular facet of medical image dissemination for radiotherapy applications that has spurred controversy, namely the anonymization of head and neck cancer (HNC) related images.

While the public dissemination of HNC image data is invaluable to improve the radiotherapy workflow, concerns have been raised regarding readily identifiable facial features on medical imaging. Importantly, the U.S. Health Insurance Portability and Accountability Act references “full-face photographs and any comparable images” as a part of protected health information (10). This policy introduces some uncertainty in the dissemination of high-resolution images, where the intricacies of facial features can be reconstructed to generate similar or “comparable” visualizations with relative ease. Several studies have shown the potential danger in releasing unaltered medical images containing facial features, as they can often be easily recognized by humans and/or machines (11–15). For example, using facial recognition software paired with image-derived facial reconstructions, one study found up to 83% of research participants could be identified from their magnetic resonance imaging (MRI) scans (13). Similar alarming results have been demonstrated for CT images (14). While brain images are often processed such that obvious facial features are removed (i.e., skull stripping), these crude techniques remove large anatomic regions necessary for building predictive models with HNC imaging data. “Defacing” tools, where voxels that correspond to the areas of the patient’s facial features are either removed or altered, offer one solution. However, they may still engender the potential loss of voxel-level information needed for predictive modeling or treatment planning, thereby prohibiting their use in data resharing strategies for radiotherapy applications. While several studies have investigated the effects of defacing for neuroimaging (16–21), there have not yet been any systematic studies on the effects of defacing tools for radiotherapy applications.

Inspired by the increasing demand for public HNC imaging datasets and the importance of protecting the privacy of patients, a systematic analysis of a number of existing methods for facial anonymization on HNC MRI images was performed. Through qualitative and quantitative analysis using open-source datasets and tools, the efficacies of defacing approaches on whole images and structures relevant to radiation treatment planning were determined. Moreover, the effects of these approaches on auto-segmentation, a specific domain application that is increasingly relevant for HNC public datasets, were also examined. This study is an important first step towards the development of robust approaches for the safe and trusted democratization of HNC imaging data.

## Methods

### Dataset

For this analysis, a publicly available dataset hosted on the TCIA, the American Association of Physicists in Medicine RT-MAC Grand Challenge 2019 (AAPM) dataset (22), was utilized. The AAPM dataset consists of T2-weighted MRI scans of 55 HNC patients that are labeled for OAR segmentations of bilateral: i) submandibular glands, ii) level II neck lymph nodes, iii) level III neck lymph nodes, and iv) parotid glands. Structures were annotated as being on the right or left side of the patient anatomy. The spatial resolution of the scans is 0.5 mm × 0.5 mm with 2.0 mm spacing. Additional technical details on the AAPM images and segmentations can be found in the corresponding data descriptor (22). Defacing experiments were also attempted using the HECKTOR 2021 training dataset (8) containing 224 HNC patients with CT scans. Additional technical details on the HECKTOR dataset can be found in the corresponding overview papers (8,9).

### Defacing methods

For defacing the images, the same methods as taken into consideration by Schwartz et al. (16), as well as novel tools that benefit from recent advances in deep learning were used. The most popular tools use a co-registration to a template in order to identify face and ears and then identify those structures in the original image, which should be removed or blurred. The following 6 co-registration based methods: *afni_refacer, fsl_deface* (23), *mask_face* (24), *mri_deface* (18), *pydeface* (25), and *quickshear* were implemented. Two more recent methods using deep learning technology were also included: *defacer* (26) and *DeepDefacer* (27). These methods utilize pre-trained deep learning models using data from public neuroimaging datasets to identify facial features to be removed. An automated pipeline for applying all these defacing methods is available at https://github.com/eglerean/faceai_testingdefacing. Each defacing method was tested with all subjects such that, for each subject, a defaced volume was produced as well as a volumetric mask of which voxels were affected by defacing. All methods were run with the default parameters and standard reference images.

### Defacing performance

After applying the defacing methods, the success or failure of a defacing method was determined by visually inspecting all the defaced volumes (i.e., performing scanwise quality control). Specifically, a binary categorization of each scan was implemented: “1” if the eyes, nose, and mouth were removed (i.e., defacing succeeded), “0” if the eyes, nose, or mouth were not removed (i.e., defacing failed). Subsequently, the amount of voxels present in the structures after application of the defacing algorithm were quantitatively measured.

### Deep learning model for OAR segmentation reliability

To evaluate the OAR segmentation performance under different defacing schemes from volumetric MRI data, a convolutional neural network architecture, 3D U-net, which has found wide success in HNC-related segmentation tasks (28–33), was utilized. Both contractive and expansive pathways include four blocks, where each block consists of two convolutional layers with a kernel size of 3, and each convolution is followed by an instance normalization layer and a LeakyReLU activation with 0.1 negative slope. The max-pooling and transpose convolutional layers have a kernel size and stride of 2. The last convolutional layer has a kernel size and stride of 1 with 9 output channels and a softmax activation. The model architecture is shown in **Figure 1**. Experiments were developed in Python v. 3.6.10 (34) using Pytorch 1.8.1 (35) with a U-net model from Project MONAI 0.7.0 (36) and data preprocessing and augmentation with TorchIO 0.18.61 (37).

**Figure 1.**
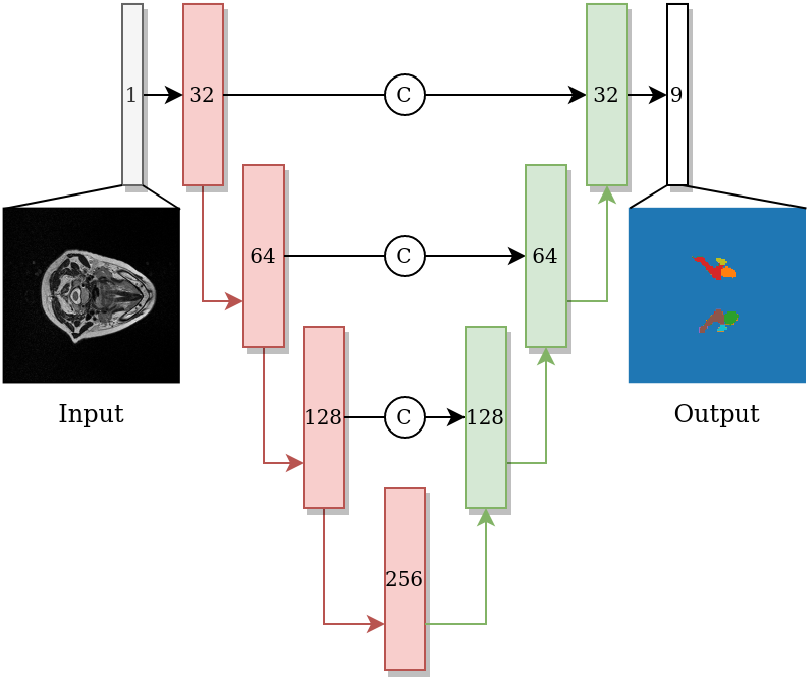
U-net network architecture with blocks on the contractive path colored in red and blocks on the expanding path colored in green. Each block includes two convolutions, each followed by instance normalization and Leaky ReLU activation, subsequently followed by a max-pool layer (red arrow) or transpose convolution layer (green arrow) on contractive and expanding paths, respectively. The number shown in each block indicates the number of channels of the feature map. Arrows with the letter C indicate concatenation.

A subset of patients for which defacing was deemed successful were used for building the segmentation models. The subset was randomly split with 5-fold cross validation: for each cross-validation iteration one fold was used for model testing, one fold was used for model validation, and the remaining three folds were used for model training. The reported segmentation performance was based on the test fold that was not used for model development. The same random splits were used for training and evaluating the models trained on original or defaced data.

Data preprocessing after the defacing included linear resampling to 2 mm isotropic resolution with the intensity scaled into a range of [-1,1]. The training data was augmented with random transforms that were applied with a probability (p), independently of each other. The used transforms were random elastic deformations (p=10%) for all axes, random flips for inferior-superior and anterior-posterior axes (p=50%), random rotation (−10° to 10°) of all axes (p=50%), random bias field (p=50%), and random gamma (p=50%). The model was trained using the cross-entropy loss for the 8 OAR classes and background with parameter updates computed using the Adam optimizer with (0.001 learning rate, 0.9 β_1_, 0.999 β_2_, and AMSGrad). The model training was stopped early after 60 epochs for non-improvement of the validation loss.

### Segmentation evaluation

Two experiments to evaluate the impact of defacing on the resulting segmentations were performed. In order to determine the impact of defacing on algorithmic development, models were trained on original or defaced data using the original target data for evaluation. Subsequently, in order to determine the impact of defacing on algorithms not originally developed for defaced data, a model was trained using the original data and its performance was evaluated by using the original data or the defaced data.

For both experiments, the performance of the models were quantified primarily with the Dice similarity coefficient (DSC) and the mean surface distance (MSD), defined as follows:

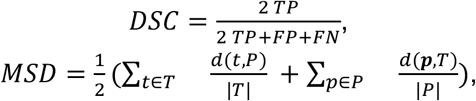

where *TP* denotes true positives, *FP* false positives, *FN* false negatives, *P* the set of segmentation surface voxels of the model output, and *T* the set of segmentation surface voxels of the annotation. The distance from the surface metric is defined as: *d(a,B) = min*_*b∈B*_{||*a −b*||_2_}. These metrics were selected because of their ubiquity in literature and ability to capture both volumetric overlap and boundary distances (38,39). The model output was resampled into the original resolution with the nearest-neighbor sampling and evaluated against the original resolution segmentations. MSD was measured in millimeters. When comparing the performance measures between the segmentation models, Wilcoxon signed rank tests (40) were implemented, with p-values less than or equal to 0.05 considered as significant. To correct for multiple hypotheses, a Benjamini-Hochberg false discovery rate procedure (41) was implemented by taking into account all the OARs and models compared. Statistical comparisons were performed using the statannotations 0.4.4 Python package (https://github.com/trevismd/statannotations). Notably, any ROI metrics that yielded empty outputs were omitted from the comparisons. Additional surface metric values (mean Hausdorff distance at 95% and Hausdorff distance at 95%) were also calculated as part of the supplementary analysis (details in **Appendix A**).

## Results

### Defacing performance

Five of the methods tested (afni_refacer, quickshear, mri_deface, DeepDefacer, and defacer) failed for all subjects in the AAPM dataset. Therefore, for all subsequent analyses only the mask_face, fsl_deface, and pydeface methods were considered. There was scanwise quality control to remove the defaced scans with poor quality from the analyses, which resulted in 16 (29%), 10 (18%), and 13 (24%) scans removed from mask_face, fsl_deface, and pydeface, respectively, with all these methods working on 29 patient scans. A barplot comparison of the ratio of remaining OAR voxels after defacing and quality control is depicted in **Figure 2**. In addition, the defacing methods removed some OARs completely, which were also omitted from the segmentation evaluation. After filtering unusable data, the total number of OARs available for use in segmentation experiments was 232 for the original data and mask_face, 231 for fsl_deface, and 169 for pydeface. A full comparison of omitted OARs is shown in **Table 1**.

**Table 1.** Quantitative details on the number of organs at risk available after the defacing was applied for all 55 patient scans. Only the mask_face, fsl_deface, and pydeface methods yielded usable data. The first group of columns correspond to the organs at risk that were completely removed from the cases with successful defacing. The second group of columns correspond to all items in the first group of columns plus incorporating any of the cases where defacing failed. Defacing success or failure was counted from scanwise quality control. *Organs at risk in these columns were omitted for all the subsequent segmentation-related experiments.

| Organ at risk / Defacing method | Completely removed after successful defacing |  |  | Unavailable for segmentation analysis* |  |  |
| --- | --- | --- | --- | --- | --- | --- |
|  | mask_face | fsl_deface | pydeface | mask_face | fsl_deface | pydeface |
| Left Submandibular Gland | 0 (0%) | 2 (4%) | 6 (11%) | 16 (29%) | 14 (25%) | 13 (24%) |
| Right Submandibular Gland | 0 (0%) | 1 (2%) | 7 (13%) | 16 (29%) | 14 (25%) | 14 (25%) |
| Left Neck Lymph Node Level II | 0 (0%) | 0 (0%) | 4 (7%) | 16 (29%) | 13 (24%) | 11 (20%) |
| Right Neck Lymph Node Level II | 0 (0%) | 0 (0%) | 4 (7%) | 16 (29%) | 13 (24%) | 11 (20%) |
| Left Neck Lymph Node Level III | 0 (0%) | 0 (0%) | 45 (82%) | 16 (29%) | 13 (24%) | 51 (93%) |
| Right Neck Lymph Node Level III | 0 (0%) | 1 (2%) | 44 (80%) | 16 (29%) | 13 (24%) | 50 (91%) |
| Left Parotid | 0 (0%) | 0 (0%) | 6 (11%) | 16 (29%) | 13 (24%) | 11 (20%) |
| Right Parotid | 0 (0%) | 0 (0%) | 6 (11%) | 16 (29%) | 13 (24%) | 11 (20%) |
| Total omitted | **0 (0%)** | **4 (1%)** | **122 (28%)** | **128 (29%)** | **106 (24%)** | **172 (39%)** |

**Figure 2.**
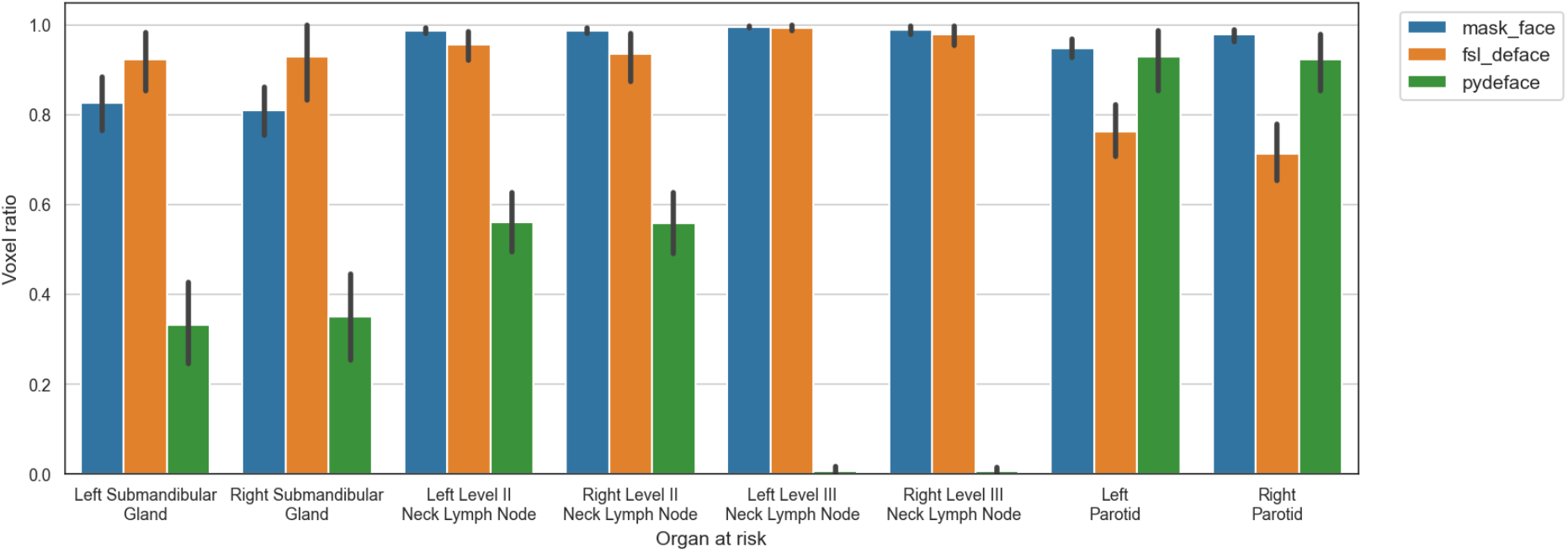
Ratio of preserved voxels in comparison to the original segmentation mask after defacing (mask_face, fsl_deface, and pydeface) for each of the organs at risk, where defacing was successful for N=39, N=42, and N=45, respectively. The mean and standard deviation are represented as the center and extremes of the error bars, respectively.

All of the tested defacing methods were unable to provide sufficient data for segmentation analysis in the HECKTOR CT dataset. Specifically, fsl_deface and pydeface methods successfully defaced 18 (8%) and 102 (46%) scans, respectively. All other methods (afni_refacer, quickshear, mri_deface, DeepDefacer, defacer, and mask_face) failed to correctly deface any of the scans. Although pydeface had the highest success rate on defacing, it only preserved the brain. Thus, no further analysis was performed for this dataset.

### Segmentation performance

The 29 patient scans for which the defacing was deemed successful were used to construct and evaluate segmentation models for the mask_face, fsl_deface, and pydeface methods. The model DSC performances pooled across all structures based on training input and valid evaluation target combinations are shown in **Table 2**. The models trained using the original, mask_face, and fsl_deface input data had the highest composite mean DSC when evaluated on the original target data with values of 0.760, 0.742, and 0.736, respectively, while the model trained on pydeface input data had the highest composite mean DSC of 0.653 when evaluated on pydeface target data. In contrast, the models trained using original mask_face, and fsl_deface input data had the lowest composite mean DSC when evaluated on pydeface target data with values of 0.406, 0.413, 0.465, respectively, while the model trained using pydeface input data had the lowest composite mean DSC of 0.395 when evaluated on fsl_deface target data. All comparisons within the same evaluation data are statistically different from each other (p≤0.05) with the exception of mask−face and fsl−deface trained models evaluated on original data. And original as well as mask−face trained models evaluated on pydeface data.

**Table 2.** Composite DSC performance-mean (standard deviation) – of all combinations of training data (rows) and evaluation data (columns). The number of total segmentation maps evaluated is shown in brackets on the header. All comparisons within the same evaluation data are statistically different from each other (p ≤ 0.05) with the exception of mask_face and fsl_deface trained models evaluated on original data, and original and mask_face trained models evaluated on pydeface data. Statistical significance was measured with Wilcoxon signed-rank tests corrected with Benjamini-Hochberg procedure comparisons within evaluation data.

|  | Evaluated on<br>original (N=232) | Evaluated on<br>mask_face<br>(N=232) | Evaluated on<br>fsl_deface<br>(N=231) | Evaluated on<br>pydeface<br>(N=169) |
| --- | --- | --- | --- | --- |
| Trained on<br>original | 0.760 (0.112) | 0.673 (0.181) | 0.693 (0.140) | 0.406 (0.304) |
| Trained on<br>mask_face | 0.742 (0.115) | 0.733 (0.120) | 0.668 (0.143) | 0.413 (0.312) |
| Trained on<br>fsl_deface | 0.736 (0.108) | 0.643 (0.185) | 0.733 (0.122) | 0.465 (0.293) |
| Trained on<br>pydeface | 0.449 (0.333) | 0.417 (0.325) | 0.395 (0.301) | 0.653 (0.258) |

### Defacing impact on model training

The analysis was based on eight OAR structure segmentations from 29 patients totaling 232 evaluations. The MSD of left and right level III neck lymph nodes for pydeface trained models were omitted from the analysis as all the model outputs were empty. Full comparisons of the model performance for each OAR are depicted in **Figure 3**. Additional surface distance metrics are shown in **Appendix A (Figure A1)**. Overall, the model trained with the original data performed better than the models trained with the defaced data for the majority of structures and evaluation metrics. Both metrics were significantly better for the model trained with the original data compared to the model trained with mask_face data for the left submandibular gland and right level II neck lymph node, while only the DSC was significantly better for the right submandibular gland and right level III neck lymph node. Similarly, both metrics were significantly better for the model trained with the original data compared to the model trained with fsl_deface data for the right level II neck lymph node, left parotid, and right parotid, while only the DSC was significantly better for the right level III neck lymph node. Moreover, both metrics were significantly better for the model trained with the original data compared to the model trained with pydeface data for all the structures.

**Figure 3.**
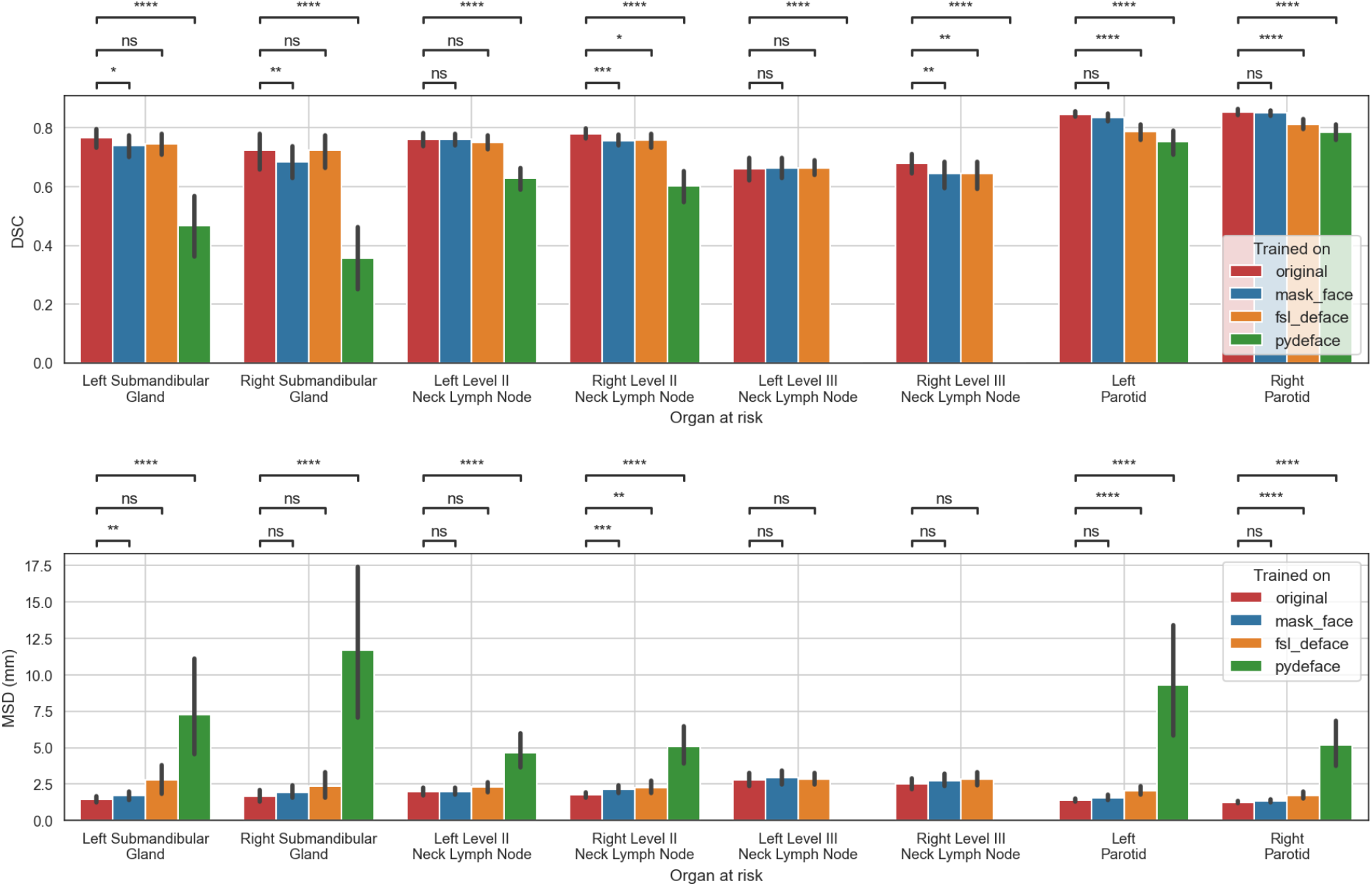
Performance of the models trained on original or defaced data and evaluated on the original data. The mean and standard deviation for each metric are represented as the center and extremes of the error bars, respectively. Statistical significance was determined using Wilcoxon signed-rank tests corrected with Benjamini-Hochberg procedure for all OARs and models. Comparison symbols: ns (p > 0.05), * (p ≤ 0.05), * * (p ≤ 0.01), * * *(p ≤ 1e-4), * * * *(p ≤ 1e-5).

### Defacing impact on model testing

In these results, only valid target data with successful defacing on all three methods using non-empty segmentation structures were included. This was obtained using results from 26 left submandibular glands, 27 right submandibular glands, 1 left neck level III lymph nodes, 2 right neck level III lymph nodes, and 28 of each of the remaining structures. Due to the low number of cases for the right and left level III lymph nodes, they were omitted from the comparison. In addition, for the MSD metric, empty model output segmentations were discarded resulting in evaluation of 1 left submandibular gland for fsl_deface and mask_face and 14 for pydeface, 1 and 6 right submandibular glands on fsl_deface and pydeface, respectively, 1 left level II lymph node for pydeface, and 2 left parotids for pydeface. The model evaluated on the original data performed significantly better than the models evaluated on the defaced data for all of the structures and both evaluation metrics except in the case of left submandibular gland DSC for fsl_deface which exhibited a non-significant difference. The full comparison of the model performance for each of the OARs is shown in **Figure 4**. Additional surface distance metrics are shown in **Appendix A (Figure A2)**.

**Figure 4.**
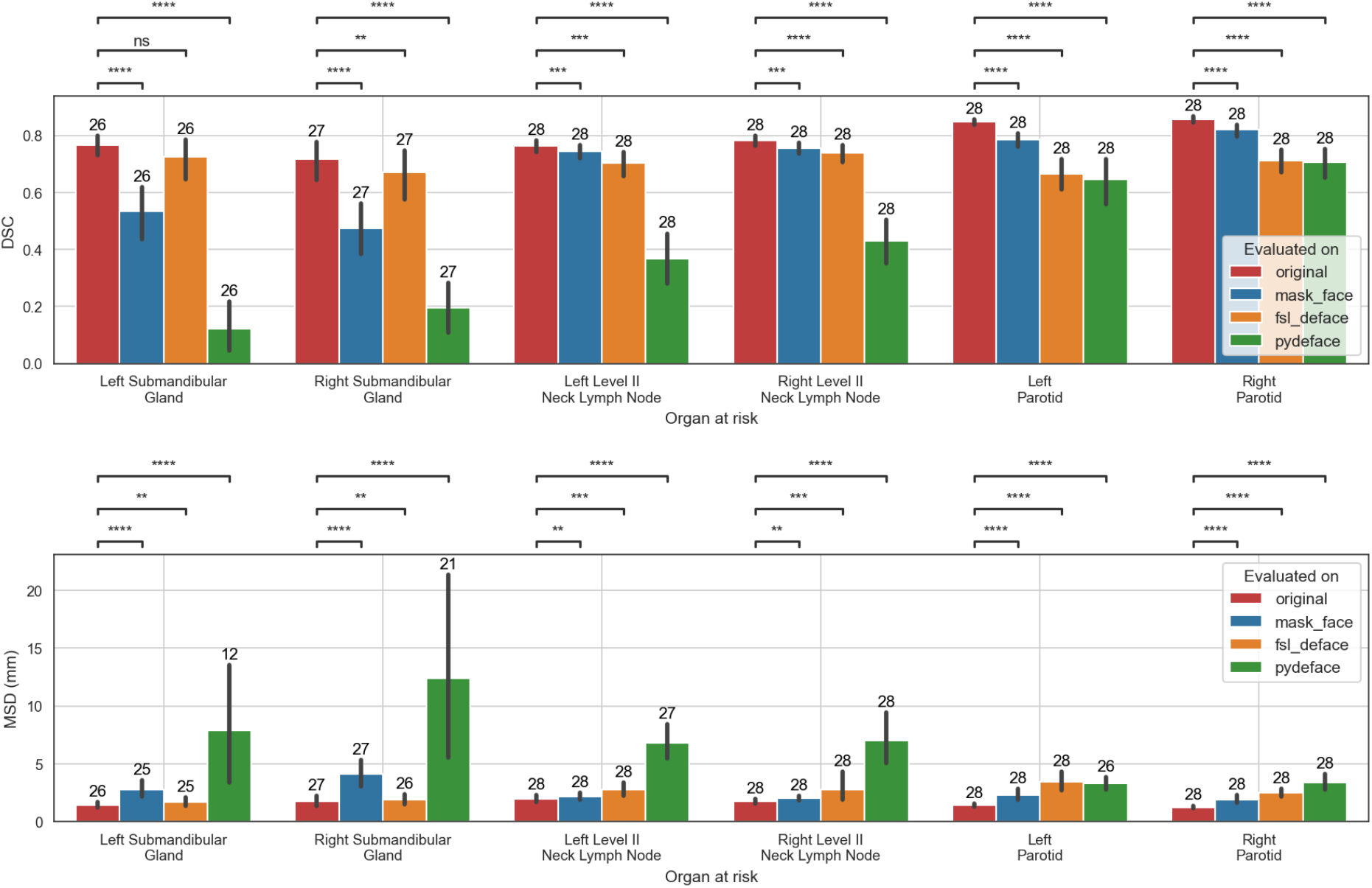
The performance of models trained on the original data when evaluated on the original, mask_face, fsl_deface, or pydeface data for the six organs at risk included in the analysis. Only cases that were available for all the methods were included: 28 segmentations were used for all structures except in the case of the left and right submandibular glands where 26 and 27 segmentations were used, respectively. In addition, for the MSD metric, empty model output segmentations were discarded, which resulted in a smaller number of evaluated structures. The number of evaluated structures is shown on top of the barplot. The mean and standard deviation for each metric are represented as the center and extremes of the error bars, respectively. Statistical significance was measured with Wilcoxon signed-rank tests corrected with Benjamini-Hochberg procedure for all OARs and models. Comparison symbols: ns (p > 0.05), * (p ≤ 0.05), * * (p ≤ 0.01), * * *(p ≤ 1e-4), * * * *(p ≤ 1e-5).

## Discussion

This study has systematically investigated the impact of a variety of defacing algorithms on structures of interest used for radiotherapy treatment planning. This study demonstrated that the overall usability of segmentations is heavily dependent on the choice of the defacing algorithm. Moreover, the results indicate that several OARs have the potential to be negatively impacted by the defacing algorithms, which is shown by the decreased performance of auto-segmentation algorithms trained and evaluated on defaced data in comparison to algorithms trained and evaluated on non-defaced data.

Defacing for HNC applications should be deemed optimal if the method simultaneously removes all recognizable facial features from the image and no voxels from structures of interest are affected. In this study, eight commonly available defacing algorithms developed by the neuroimaging community were applied: afni_refacer, mri_deface, defacer, DeepDefacer, mask_face, fsl_deface, pydeface, and quickshear. Unfortunately, for the investigated CT data, no defacing method was able to yield successful removal of facial features while preserving the OARs. This is not necessarily surprising given that the methods investigated were developed primarily with MRI in mind; these results echo previous similar work using CT data (42). Importantly, even when applied to MRI data of HNC patients, many of these defacing methods outright failed for most if not all patients. Therefore, despite extant studies demonstrating the acceptability of these methods to remove facial features from neuroimaging scans (16–21), these tools may not necessarily be robust to HNC-related imaging. Moreover, for those defacing algorithms that were able to successfully remove facial information in the MRI data, i.e. mask_face, fsl_deface, and pydeface, it was shown that regardless of the choice of the method, there was a loss of voxel-level information for all the OAR structures investigated. Importantly, pydeface leads to a greater number of lost voxels than mask_face and fsl_deface for all the OAR structures, with the exception of the parotid glands. While mask_face and fsl_deface lead to relatively minimal reduction of available voxels in many cases, the loss of topographic information in a radiotherapy workflow cannot be underscored enough. It is well known that even minor variations in the delineation of tumors and OARs can drastically alter the resulting radiotherapy dose delivered to a patient, which can impact important clinical outcomes such as toxicity and overall survival (43–46). Therefore, the loss of voxel-level information of OARs caused by the defacing algorithms, while potentially visibly imperceptible, can still affect downstream clinical workflows.

Relatively few studies have been conducted that determined the downstream analysis effects of defacing algorithms. For example, recent studies by Schwartz et al. (16) and Mikulan et al. (21) demonstrated that several defacing methods showed differences in specific neuroimaging applications, namely brain volume measurements and electroencephalography-related calculations. In this study, as a proxy for a clinically relevant task, an OAR auto-segmentation workflow was developed to investigate the impact of defacing-induced voxel-level information loss on downstream radiotherapy applications. As evident through both pooled analysis and investigation of individual OARs for auto-segmentation model training and evaluation, performance is often modestly decreased for fsl_deface and mask_face but greatly decreased for pydeface; these results were consistent with the overall voxel-level information loss. While pydeface has been shown to have favorable results for use with neuroimaging data (19,21), its negative impact on HNC imaging is apparent. Therefore, in cases where defacing is unavoidable, mask_face or fsl_deface should likely be preferred for HNC image anonymization. Regardless, this study demonstrates existing approaches to anonymize facial data may not be sufficient for implementation on HNC-related datasets, particularly for deep learning model training and testing.

This study has several limitations. Firstly, to examine defacing methods as they are currently distributed (“out-of-the-box”), modifications to the templates or models utilized in any methods were not performed. Further preprocessing either of the CT and MRI data as well as subject specific settings could have helped some of the methods to better identify the face. In addition, more suitable templates for the HNC images (for both CT and MRI) would likely improve the defacing performance; for the registration-based algorithms, algorithms likely expected scans to cover the whole brain, while the field-of-view of the images for HNC mostly covered the neck and mouth, leaving the top of the brain excluded. Notably, additional deep learning model training schemes (i.e., transfer learning) may potentially allow for eventual implementation of existing deep learning methods on domain-specific datasets (i.e., HNC radiotherapy), but this negates the immediate interoperability of these tools. Furthermore, no additional image processing other than what was integrated into the defacing methods was implemented; it may be possible alternative processing could alter these results. Secondly, while a robust analysis utilizing multiple relevant metrics established in existing literature (38) was performed to evaluate OAR auto-segmentation, there is not always a perfect correlation between spatial similarity metrics and radiotherapy plan acceptability (39). This study has not tested the downstream effects of defacing on radiotherapy plan generation, which may lead to different results from what was observed for the OAR segmentation. Thirdly, this study was limited to public data with no modifications. Only structures that were already available in existing datasets were analyzed. Moreover, as an initial exploration of defacing methods for radiotherapy applications, only a single imaging modality on a relatively limited sample size, namely T2-weighted MRI, was investigated for auto-segmentation experiments, despite the HNC radiotherapy workflow commonly incorporating additional modalities (47). Thus, experiments on additional imaging modalities and larger diverse HNC patient populations should be the subject of future investigations. Fourthly, the current analysis does not thoroughly explore possible performance confounding related to phenotypical and individual variables such as sex, ethnicity, and age of the measured individuals. Finally, this study has focused on defacing methods as an avenue for public data sharing for training and evaluating machine learning models, but privacy-preserving modeling approaches, e.g., through federated learning (48), may also act as a potential alternative solution.

## Conclusions

In summary, by using publicly available data, the effects of eight established defacing algorithms, afni_refacer, mask_face, mri_deface, defacer, DeepDefacer, quickshear, fsl_deface, and pydeface, have been systematically investigated for radiotherapy applications. Specifically, the impact of defacing directly on ground-truth HNC OARs was determined and a deep learning based OAR auto-segmentation workflow to investigate the use of defaced data for algorithmic training and evaluation was developed. All methods failed to properly remove facial features on the CT dataset investigated. Moreover, it was observed that only fsl_deface, mask_face, and pydeface yielded usable images from the MRI dataset, but still decreased the total number of voxels in OARs and negatively impacted the performance of OAR auto-segmentation, with pydeface having more severe negative effects than mask_face or fsl_deface. This study is an important step towards ensuring widespread privacy-preserving dissemination of HNC imaging data without endangering data usability. Given that current defacing methods remove critical data, future larger studies should investigate alternative approaches for anonymizing facial data that preserve radiotherapy-related structures. Moreover, studies on the impact of these methods on radiotherapy plan generation, the inclusion of a greater number of OARs and target structures, and the incorporation of additional imaging modalities are also warranted.

## Data Availability

All data produced in the present study are available upon reasonable request to the authors.

## Funding Acknowledgements

This work was supported by the National Institutes of Health (NIH)/National Cancer Institute (NCI) through a Cancer Center Support Grant (CCSG; P30CA016672-44). M.A.N. is supported by an NIH grant (R01DE028290-01). K.A.W. is supported by a training fellowship from The University of Texas Health Science Center at Houston Center for Clinical and Translational Sciences TL1 Program (TL1TR003169), the American Legion Auxiliary Fellowship in Cancer Research, and an NIH/National Institute for Dental and Craniofacial Research (NIDCR) F31 fellowship (1 F31DE031502-01). C.D.F. received funding from the NIH/NIDCR (1R01DE025248-01/R56DE025248); an NIH/NIDCR Academic-Industrial Partnership Award (R01DE028290); the National Science Foundation (NSF), Division of Mathematical Sciences, Joint NIH/NSF Initiative on Quantitative Approaches to Biomedical Big Data (QuBBD) Grant (NSF 1557679); the NIH Big Data to Knowledge (BD2K) Program of the NCI Early Stage Development of Technologies in Biomedical Computing, Informatics, and Big Data Science Award (1R01CA214825); the NCI Early Phase Clinical Trials in Imaging and Image-Guided Interventions Program (1R01CA218148); an NIH/NCI Pilot Research Program Award from the UT MD Anderson CCSG Radiation Oncology and Cancer Imaging Program (P30CA016672); an NIH/NCI Head and Neck Specialized Programs of Research Excellence (SPORE) Developmental Research Program Award (P50CA097007); and the National Institute of Biomedical Imaging and Bioengineering (NIBIB) Research Education Program (R25EB025787).

## Author contributions

Study concepts: all authors; Study design: J.S., E.G., J.J; Data acquisition: K.A.W., M.A.N., R.H; Quality control of data and algorithms: J.S., E.G.; Data analysis and interpretation: J.S., K.A.W., E.G., J.J., B.H.K., A.M., K.K; Manuscript editing: J.S., K.A.W., E.G., J.J., B.H.K., A.M., K.K, C.D.F. All authors contributed to the article and approved the submitted version.

## Appendix A: Supplementary Data

For completeness, segmentation experiments were also quantified using additional surface distance metrics. These metrics were the mean Hausdorff distance at 95% (MHD_95_) and the Hausdorff distance at 95% (95HD):

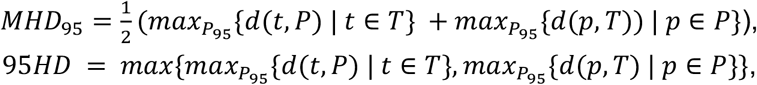

where *P* the set of segmentation surface voxels of the model output, and *T* the set of segmentation surface voxels of the annotation. The distance from the surface metric is defined as: *d(a,B) = min*_*b∈B*_{||*a−b*||_2_}.

Additional metrics for the model training and model testing experiments are shown in **Figure A1** and **Figure A2**, respectively.

**Figure A1.**
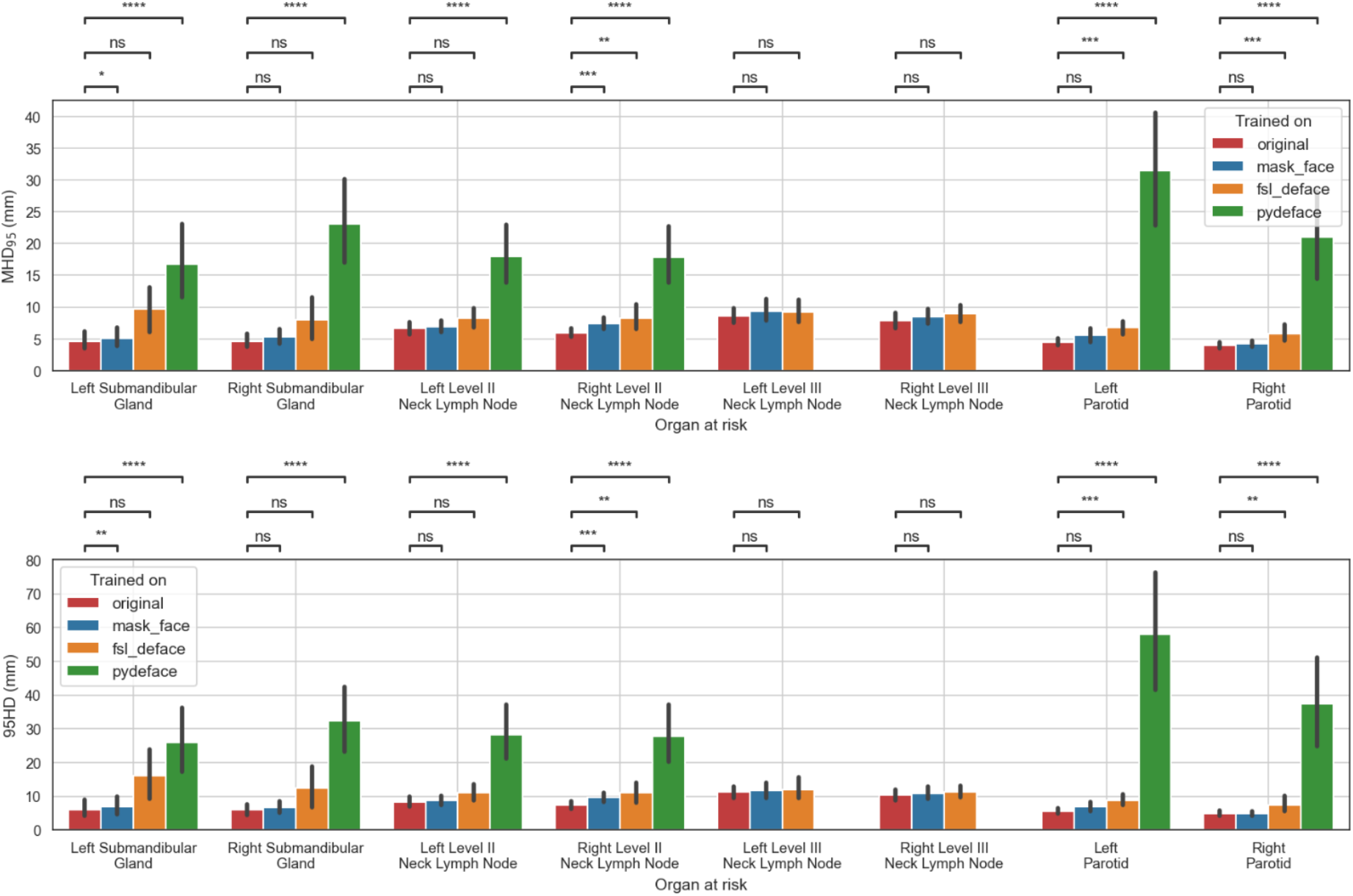
Additional surface metric values for performance of models trained on the original or defaced data and evaluated on the original data. The mean and standard deviation for each metric are represented as the center and extremes the error bars, respectively. Statistical significane was determined using Wilcoxon signed-rank tests corrected with Benjamini-Hochberg procedure for all OARS and models. Comparison symbols: ns(p > 0.05), * (p ≤ 0.05), * * (p ≤ 0.01), * * *(p ≤ 1e-4), * * * * (p ≤ 1e-5).

**Figure A2.**
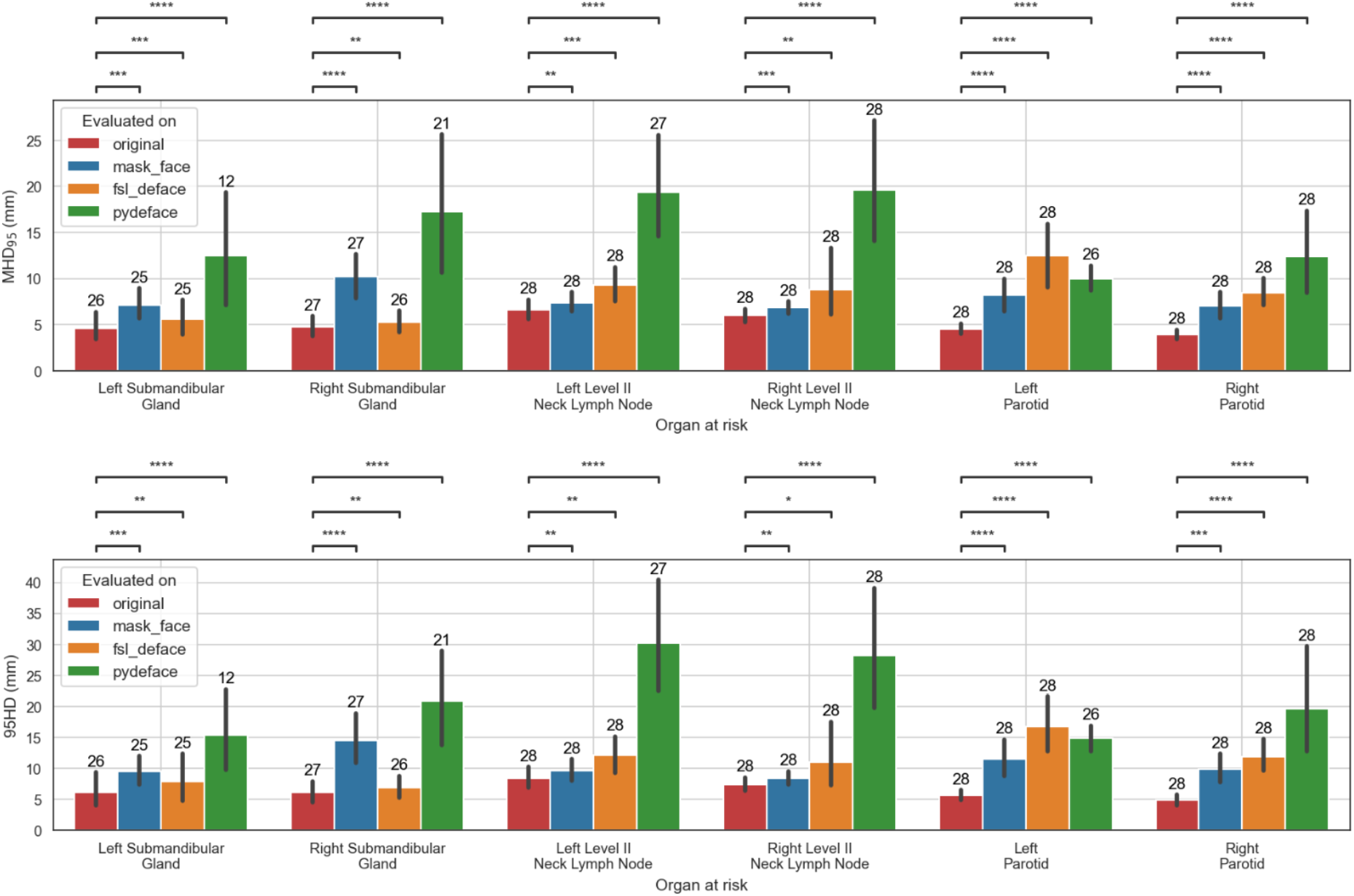
Additional surface metric values for performance of models trained on the original data when evaluated on the original, mask_face, fsl_deface, or pydeface data for the six organs at risk included in the analysis. Only cases that were available for all methods were included: 28 segmentations were used for all structures except in the case of the left and right submandibular glands where 26 and 27 segmentations were used, respectively. Empty model output segmentations were discarded, which resulted in a smaller number of evaluated structures. The number of evaluated structures is shown on top of the barplot. The mean and standard deviation for each metric are represented as the center and extremes of the error bars, respectively. Statistical significance was measured with Wilcoxon signed-rank tests corrected with Benjamini-Hochberg procedure for all OARs and models. Comparison symbols: ns (p >0.05), * (p ≤ 0.05), * * (p ≤ 0.01), * * *(p ≤ 1e-4), * * * * (p ≤ 1e-5).

## Notes

### Competing Interest Statement

C.D.F. has received direct industry grant support, speaking honoraria, and travel funding from Elekta AB. The other authors have no conflicts of interest to disclose.

### Funding Statement

This work is supported by the International Laboratory of Social Neurobiology ICN HSE RF Government grant ag. No. 075 15 2019 1930 (to E.G.). This work was also supported by the National Institutes of Health (NIH)/National Cancer Institute (NCI) through a Cancer Center Support Grant (CCSG; P30CA016672-44). M.A.N. is supported by an NIH grant (R01DE028290-01). K.A.W. is supported by a training fellowship from The University of Texas Health Science Center at Houston Center for Clinical and Translational Sciences TL1 Program (TL1TR003169), the American Legion Auxiliary Fellowship in Cancer Research, and an NIH/National Institute for Dental and Craniofacial Research (NIDCR) F31 fellowship (1 F31DE031502-01). C.D.F. received funding from the NIH/NIDCR (1R01DE025248-01/R56DE025248); an NIH/NIDCR Academic-Industrial Partnership Award (R01DE028290); the National Science Foundation (NSF), Division of Mathematical Sciences, Joint NIH/NSF Initiative on Quantitative Approaches to Biomedical Big Data (QuBBD) Grant (NSF 1557679); the NIH Big Data to Knowledge (BD2K) Program of the NCI Early Stage Development of Technologies in Biomedical Computing, Informatics, and Big Data Science Award (1R01CA214825); the NCI Early Phase Clinical Trials in Imaging and Image-Guided Interventions Program (1R01CA218148); an NIH/NCI Pilot Research Program Award from the UT MD Anderson CCSG Radiation Oncology and Cancer Imaging Program (P30CA016672); an NIH/NCI Head and Neck Specialized Programs of Research Excellence (SPORE) Developmental Research Program Award (P50CA097007); and the National Institute of Biomedical Imaging and Bioengineering (NIBIB) Research Education Program (R25EB025787).

### Author Declarations

The study involves only openly avalailbe human data, which can be obtained from: https://wiki.cancerimagingarchive.net/display/Public/AAPM+RT-MAC+Grand+Challenge+2019. As per the corresponding description of this dataset: "Images of patients were retrospectively retrieved under an Institutional Review Board approved protocol and waiver of informed consent (RCR08-0300).”

### Summary of Updates

Several sections have been restructured and new data has been added.

